# Potential reduction in transmission of COVID-19 by digital contact tracing systems: a modelling study

**DOI:** 10.1101/2020.08.27.20068346

**Authors:** Michael J. Plank, Alex James, Audrey Lustig, Nicholas Steyn, Rachelle N Binny, Shaun C. Hendy

## Abstract

**Background:** Digital tools are being developed to support contact tracing as part of the global effort to control the spread of COVID-19. These include smartphone apps, Bluetooth-based proximity detection, location tracking, and automatic exposure notification features. Evidence on the effectiveness of alternative approaches to digital contact tracing is so far limited.

**Methods:** We use an age-structured branching process model of the transmission of COVID-19 in different settings to estimate the potential of manual contact tracing and digital tracing systems to help control the epidemic. We investigate the effect of the uptake rate and proportion of contacts recorded by the digital system on key model outputs: the effective reproduction number, the mean outbreak size after 30 days, and the probability of elimination.

**Results:** Effective manual contact tracing can reduce the effective reproduction number from 2.4 to around 1.5. The addition of a digital tracing system with a high uptake rate over 75% could further reduce the effective reproduction number to around 1.1. Fully automated digital tracing without manual contact tracing is predicted to be much less effective.

**Conclusions:** For digital tracing systems to make a significant contribution to the control of COVID-19, they need be designed in close conjunction with public health agencies to support and complement manual contact tracing by trained professionals.

## Introduction

Contact tracing has been crucial in controlling several disease outbreaks, notably SARS, MERS and Ebola (WHO & CDC, 2015; Kang et al., 2015) and has become a key tool in the global effort to control the spread of COVID-19. While contact tracing alone is unlikely to contain the spread of COVID-19 (Hellewell et al., 2020; Kucharski et al., 2020), in countries where cases have been reduced to very low numbers, it may allow population-wide social distancing measures to be relaxed. In countries with more widespread epidemics, it can allow safe reopening.

Manual contact tracing typically involves interviewing confirmed cases about their recent contacts, and asking those contacts to take measures to prevent onward transmission of the disease in case they are infected. Such measures may include limiting their interactions with others, formal quarantine, getting tested, or remaining vigilant for symptoms. Manual contact tracing is intensive work and requires highly trained public health professionals to be implemented effectively (Verrall, 2020). It is also difficult to scale manual contact tracing up to deal with very large outbreaks.

In response, many countries have attempted to develop digital contact tracing systems using smartphone apps. There are multiple different approaches to this problem, for example QR code-based systems, Bluetooth-proximity based apps, automated exposure notification features, and systems that do not rely on smartphones such as card-based proximity detection (Anglemyer et al., 2020). Digital systems can offer, to varying extents, three main benefits to controlling COVID-19: (i) an increase in the proportion of contacts who are traced (e.g. contacts that would not be traced by case recall but are recorded by the digital system); (ii) a reduction in the time taken to identify and notify traced contacts (e.g. via an exposure notification feature); (iii) improved scalability over manual contact tracing.

The effectiveness of digital contact tracing is still unproven, with limited real-world data (Anglemyer et al., 2020). Mathematical modelling is useful because it can estimate the effect of digital tracing systems under different assumptions about their uptake and effectiveness (Ferretti et al., 2020; Kucharski et al., 2020). Here, we use a model of COVID-19 transmission and contact tracing to evaluate the potential of digital contact tracing systems to reduce the spread of COVID-19. We evaluate the benefits of digital contact tracing both alone and in combination with manual tracing, over a range of uptake rates, tracing probabilities, and the effectiveness of quarantine.

## Methods

### Transmission model

We use an age-structured branching process model for COVID-19 transmission and contact tracing that is an extension of the age-structured model of James et al. (2021a) to include different contact types: home, work, school and casual. These contact types are assumed to have secondary attack rates of 20% for home contacts and 6% for work, school and casual contacts (Kucharski et al., 2020), independent of age (Bi et al., 2020). Age-specific contact patterns in each of these four settings are based on the contact matrices estimated by Prem at al. (2017) for New Zealand (see Supplementary Figure S1), although contact patterns are similar for other comparable countries. The contact rates were scaled by a parameter to give a basic reproduction number in the absence of any case-targeted control of *R*_0_ = 2.6 (Jarvis et al., 2020; Kucharski et al., 2020).

The time from infection to symptom onset is gamma distributed with mean 5.5 days and standard deviation 2.3 days (Lauer et al., 2020). The generation interval is assumed to follow a Weibull distribution with mean 5.1 days and standard deviation 1.9 days (Ferretti et al., 2020), time-shifted such that 35% of transmission occurs prior to symptom onset (Ganyani et al., 2020). Infections have an age-dependent probability of being subclinical (Davies et al., 2020) that is assumed to decrease linearly from 40% in the 0-10 year age group to 5% in the over 70 years age group. Subclinical individuals are assumed to be 50% as infectious as clinical individuals.

We model individual heterogeneity and homophily in number of contacts. Heterogeneity is modelled by individual multipliers 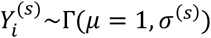 for the number of contacts that individual *i* has in setting s. The parameter *σ*^(*s*)^ represents the level of heterogeneity in individual contact rates in setting *s*, with higher values of *σ*^(*s*)^ corresponding to a greater degree of superspreading in the transmission dynamics (Lloyd-Smith et al., 2005). We assume *σ*^(*s*)^ is higher for work, school and casual contacts than for home contacts (see Table 1), reflecting lower opportunity for superspreading events in the home setting.

**Table 1.** Model parameters values and references.

| Parameter | Value | Source |
| --- | --- | --- |
| Secondary attack rate | $A^{(home)} = 20\%$ | Kucharski (2020) |
| | $A^{(work,school,casual)} = 6\%$ | |
| Basic reproduction number | $R_0 = 2.6$ | Kucharski (2020)<br>Jarvis (2020) |
| Variance in individual contact rates | $\sigma^2 (home) = 1$ | Riou (2020) |
| | $\sigma^2 (work,school,casual) = 2$ | James (2021b) |
| Probability of infections in age group $k$ (in 10-year age bands) being subclinical | $p_{sub}(k) = [0.40, 0.35, \dots, 0.10, 0.05]$ | Davies (2020) |
| Relative infectiousness of subclinical individuals | $c_{sub} = 50\%$ | Davies (2020) |
| Incubation period (days) | $T_{onset} \sim \Gamma(\text{shape} = 5.8, \text{scale} = 0.95)$ | Lauer (2020) |
| Generation time distribution (days) | $T_{transmit} - T_{onset} + t_s \sim Weibull(\text{shape} = 2.83, \text{scale} = 5.67)$ | Ferretti (2020) |
| Probability of testing (clinical individuals) | $p_{detect} = 1$ | |
| Time from onset to isolation and testing (untraced contacts) (days) | $T_{iso} \sim \Gamma(\text{shape} = 1.3, \text{scale} = 5.2)$ | James (2021b) |
| Distribution of testing to test result (days) | $T_{result} - 0.5 \sim Exp(0.5)$ | James (2021a) |
| Proportion of pre-symptomatic transmission | $p_{pre} = 35\%$ | Ganyani (2020)<br>Tindale (2020) |
| Relative infectiousness after quarantine | $c_{quar} = 50\%, 20\%$ | |
| Relative infectiousness after isolation | $c_{iso} = 0$ | |
| Homophily parameter | $q = 0.5$ | |
| Manual tracing probability | $P_{manual} = \begin{cases} 1 & \text{home} \\ 0.8 & \text{school} \\ 0.5 & \text{work} \\ 0.25 & \text{casual} \end{cases}$ | Kucharski (2020) |
| Time from test result to contact tracing for manually traced contacts (days) | $T_{trace} = \begin{cases} 0 & \text{home} \\ 0.5 & \text{school} \\ \Gamma(\mu = 3, \sigma = 1.7) & \text{work} \\ \Gamma(\mu = 3, \sigma = 1.7) & \text{casual} \end{cases}$ | Ferretti (2020)<br>James (2021a) |
| Digital tracing probability (provided index case and contact are both users) | $P_d = \begin{cases} - & \text{home} \\ 0.9 & \text{school} \\ 0.9 & \text{work} \\ 0.9 & \text{casual} \end{cases}$ | |

We model homophily by assuming that, when individual *i* infects individual *j* in setting *s* = *s*\*(work, school or casual) 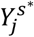 is equal to 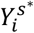 with probability *q*, and is independently sampled from 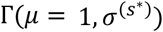 with probability 1 − *q*. This models a situation where people with high contact rates in setting s tend to be linked to other people with high contact rates in setting *s* and vice versa, where *q* is a parameter measuring the level of homophily in contact patterns. Values of 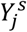 for different settings (*s* ≠ *s*\*) are assumed to be independently sampled from Γ(*μ* = 1, *σ*^(*s*)^).

Home transmissions are treated differently to model spread of the virus among a household containing a fixed number of members. Each infected individual is assigned a household identification number and an expected number of home contacts, given by 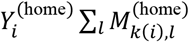, where 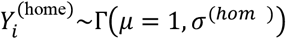 When a secondary infection *j* occurs via household transmission, the expected number of household contacts for the newly infected individual decreases by one with each new infection in the same household. This means that, eventually, transmission chains within a given household will always go extinct as the pool of susceptible household members is depleted. For simplicity, network effects and infection-induced immunity for work, school and casual contacts are ignored, i.e. all non-household contacts of the secondary infection are assumed to be mutually exclusive of the non-household contacts of the index infection.

The number of people infected by individual *i* in setting s and age group *l* between time *t* and time *t* + at is *δt* a Poisson random variable with mean:

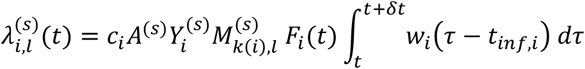

where:

- *c*_*i*_ = 0.5 if individual *i* is subclinical and *c*_*i*_ = 1 if individual *i* is clinical.
- *A*^(*s*)^ is the secondary attack rate for contacts in setting *s*.
- 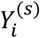 models individual heterogeneity in transmission rates for setting *s*.
- 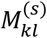 is the contact matrix representing the average number of contacts that an individual in age group *k* has in age group *l* and setting *s*.
- *k*(*i*) is the age group of individual *i*.
- *F*_*i*_(*t*) is equal to *c*_*quar*_ if individual *i* is in quarantine at time *t, c*_*iso*_ if individual *i* is in isolation at time *t*, or 1 otherwise (see *Testing and contact tracing model* below).
- *t*_*inf,i*_ is the time individual *i* was infected.
- *W*_*i*_ is the probability density function of the generation time distribution for individual *i*. This is defined to be 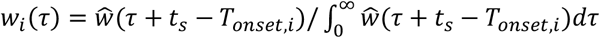, where *ŵ* is a Weibull distribution with mean 5.1 days, s.d. 1.9 days, *T*_*onset,i*_ is the symptom onset time for individual *i*, and *t*_*s*_ = 4.2 days is a constant that shifts the generation time distribution such that, in the absence of any case isolation measures, 35% of transmission occurs before symptom onset.

A complete list of parameter values is shown in Table 1. Supplementary Figure S2 shows an example simulation of the branching process and contact tracing model.

### Testing and contact tracing model

In the absence of any contact tracing, clinical individuals are assumed to be tested eventually with probability *p*_*detect*_ = 1 (i.e. all clinical infections are eventually detected). The delay from onset of symptoms to testing is assumed to be gamma distributed with mean 6.8 days and standard deviation 5.9 days (James et al., 2021b). Cases are isolated at the same time as getting tested and this prevents any further transmission. There is a further delay between getting tested and the test result being returned that is a minimum of 0.5 days, plus an exponentially distributed random variable with a mean of 0.5 days. Subclinical individuals do not get tested and do not isolate.

When a new positive test result is returned and contact tracing for that case begins, we refer to the individual testing positive as the index case and to infected contacts of the index case as secondary cases. Tracing of the index case’s contacts begins when the index case returns a positive test result (see Figure 1). Under manual tracing, each contact has a setting-dependent probability of being traced and time taken to trace. Traced contacts who are not currently symptomatic (i.e. either subclinical or pre-symptomatic) are quarantined, which is assumed to reduce onward transmission to a level *c*_*quar*_ < 1 relative to no quarantine. We investigate two scenarios where quarantine reduces transmission by 50% (*c*_*quar*_ = 0.5) or 80% (*c*_*quar*_ = 0.2). Traced contacts who become symptomatic go into isolation immediately on symptom onset, which is assumed to completely prevent any further onward transmission. Effective contact tracing therefore reduces transmission in two ways: (i) quarantining of contacts who are not currently symptomatic; and (ii) prompt isolation of secondary cases immediately on symptom onset.

**Figure 1.**
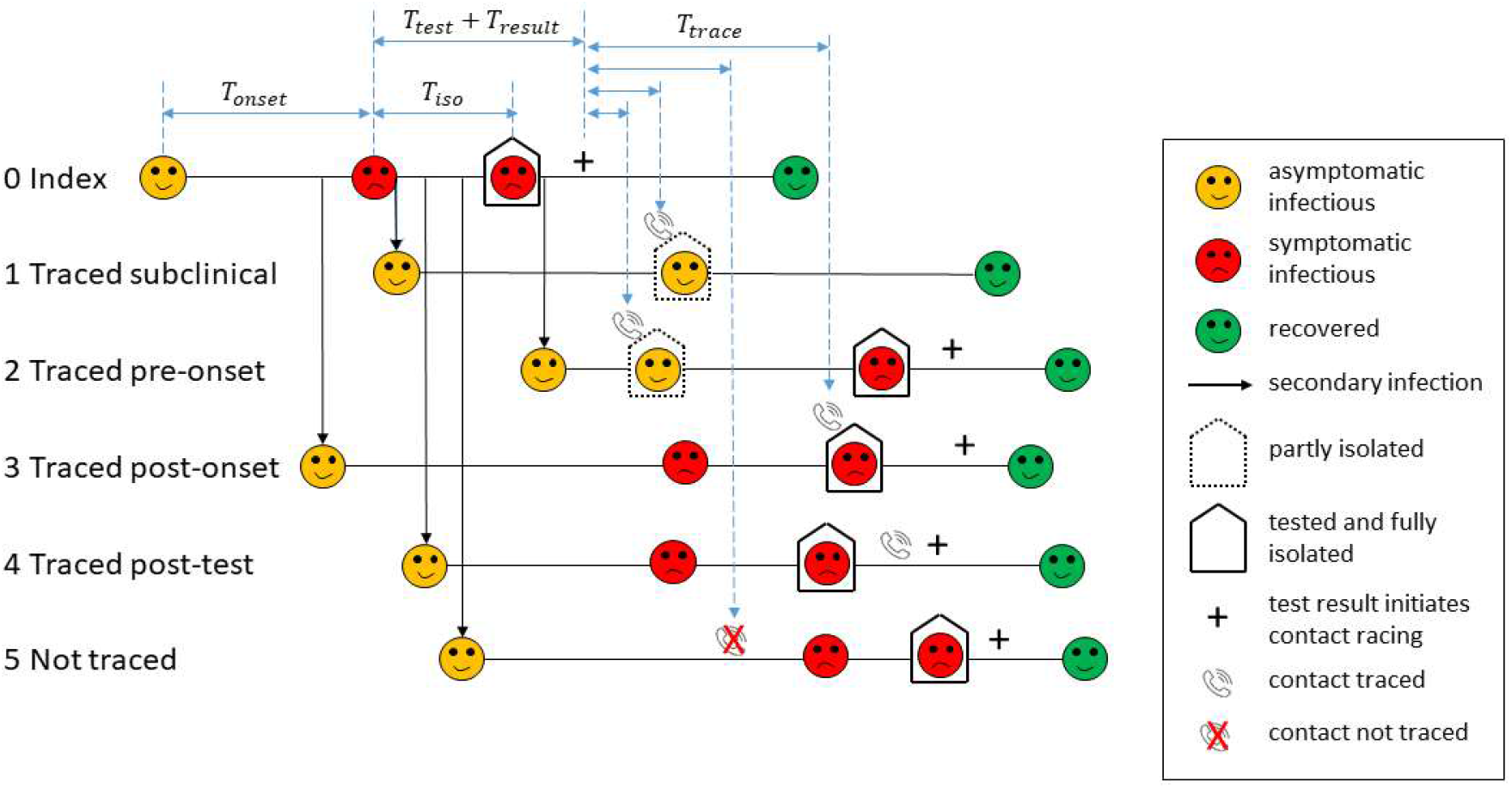
Schematic diagram of the contact tracing model. Infectious individuals are initially asymptomatic (yellow). For the index case who was not traced (0), there is a delay between onset of symptoms (red) and getting tested. Isolation occurs at the same time as getting tested. There is a subsequent delay to the test result being returned (+) and tracing of contacts. Traced contacts (1-4) are quarantined when contacted by public health officials (phone icons) and are isolated and tested immediately on symptom onset. Traced contacts (3) who are already symptomatic prior to being traced are isolated immediately when contacted. Traced contacts (4) that have already isolated prior to being traced are not affected. Contacts that cannot be traced (5) may still get tested and isolated, but this is likely to take longer. Subclinical individuals (1) do not get tested or isolated, but will be quarantined if they are a traced contact.

Home contacts are assumed to be traced instantly (i.e. immediately after the index case returns a positive test result) with probability 1, independently of any digital contact tracing system. Under manual contact tracing, work contacts are traced with probability 0.5, school contacts with probability 0.8, and casual contacts with probability 0.25. These are similar to the tracing probabilities for different contact types estimated by Kucharski et al. (2020). Work and casual contacts are assumed to have a tracing time that is gamma distributed with mean 3 days and standard deviation 1.7 days (Ferretti et al., 2020; James et al., 2021a). School contacts are assumed to be traced more rapidly but not instantly (0.5 days after the index case returns a positive test result).

### Digital contact tracing systems

We model digital contact tracing systems by varying key parameters of the contact tracing model. Home contacts are assumed to be always traced rapidly by the manual system, so digital tracing only applies to school, work and casual contacts. For each scenario, we assume there is an uptake rate *u* and that each individual in the population is a user of the digital tracing system (e.g. has installed the app) with probability *u*, independently of all other individuals. This ignores any correlation in the usage probabilities of close contacts.

Provided the index case and secondary case are both users of the system, the contact is digitally logged with probability *P*_*d*_, which we assume is the same for school, work and casual contacts (see Table 1). We set a default value of *P*_*d*_ = 0.9, but also investigate values of P_d_ smaller than this. A tracing probability of 100% is likely to be unachievable, for example because there will be situations where one or both individuals do not carry the smartphone or card with them. Contacts that are digitally logged are assumed to be quarantined immediately after the index case returns a positive test result, the same as for home contacts. If neither or one of the index case and the secondary case is a user of the system, or both are users but the contact was not logged by the digital system, the contact is not traced digitally, but may still be later traced manually.

We investigate how the performance of the contact tracing system varies with the uptake rate *u* and the probability of digital tracing *P*_*d*_ (see Table 1). We assume the effectiveness of quarantine is the same for digitally and manually traced contacts. This is likely to require effective manual follow up of digitally traced contacts as opposed to relying solely on automatic exposure notifications (James et al., 2021a).

We also investigate the additional benefit from including recursive tracing of second-order contacts (i.e. quarantining the contacts of contacts of a confirmed case) in the model. Recursive tracing and effective quarantine of second-order contacts is more difficult to achieve in practice because of the much larger number of second-order contacts and the lower risk of them being infected. In the case on an ongoing outbreak with a large number of cases, the number of uninfected individuals being quarantined under recursive tracing is likely to be prohibitively large (Firth et al., 2020). However, recursive tracing could potentially be useful in suppressing a small outbreak in its very early stages. We assume that tracing of second-order contacts begins 2 days after tracing of the first-order contact and can occur digitally or manually following the same rules described above (Figure 2). This means that second-order contacts who are traced digitally are quarantined 2 days after quarantining the first-order contact; second-order contacts traced manually are quarantined later. Any second-order contacts made subsequent to quarantine of the first-order contact cannot be traced recursively, but may still be traced on or after the positive test result of the first-order contact. Third-order contacts were not traced recursively.

**Figure 2.**
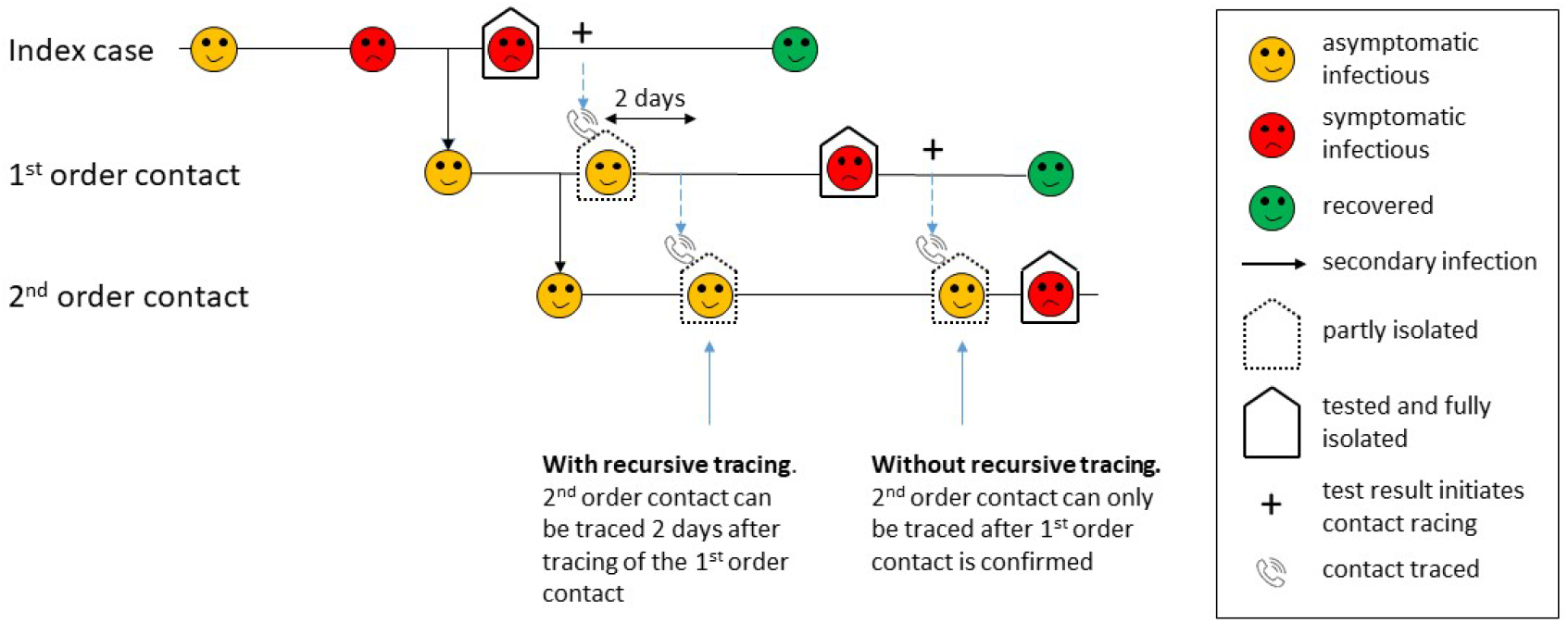
Recursive tracing of second-order contacts. With recursive tracing, second-order contacts of a confirmed case can be traced and quarantined two days after the first-order case is traced and quarantined (assuming both contacts are traced digitally).

## Results

We measured the reduction in spread of COVID-19 by looking at three model outputs: (a) the effective reproduction number *R*_*eff*_ (average number of secondary infections per case); (b) the mean outbreak size (total number of infections per seed case) after 30 days; (c) the probability of extinction of an outbreak starting from a single seed case. Together these outputs measure the relative effectiveness of the contact tracing system in containing the virus. Results are robust to the initial number of seed cases: if there are multiple initial seed cases, the mean reproduction number is not affected; the mean outbreak size is linear in the initial number of cases, and the probability of elimination is raised to the power of the initial number of cases. For each combination of contact tracing parameters, we ran multiple simulations, each initialised with a single infected seed case. Results are shown for uptake rates ranging from 0 to 90% and were calculated by averaging over 10,000 independent realisations of the branching process, each run until the outbreak was either eliminated or reached 1,000 cumulative cases.

With case isolation in the absence of any contact tracing, the effective reproduction number was *R*_*eff*_ = 2.4, the mean outbreak size after 30 days was 78, and the probability of extinction was approximately 47%. Manual-only contact tracing (which corresponds to a digital uptake rate of *u* = 0 in Fig. 3) with moderately effective (reduces transmission by 50%) quarantine of pre-symptomatic or subclinical individuals reduced *R*_*eff*_ to 1.56, the mean outbreak size to approximately 36 and increased the probability of extinction to 63%.

**Figure 3:**
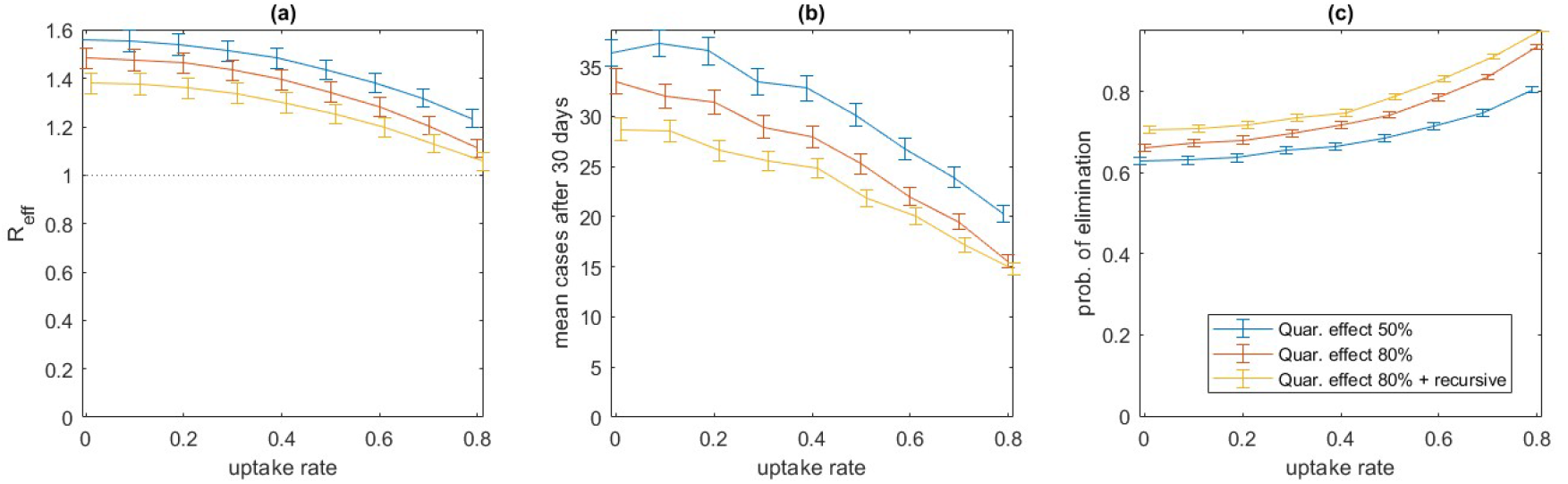
Effectiveness of manual contact tracing plus digital tracing at a range of uptake rates, under different effectiveness of quarantine, with and without recursive tracing. Effectiveness of manual contact tracing supported by a digital tracing system measured by: (a) effective reproduction number *R*_*eff*_ (mean number of secondary infections per case); (b) mean total number of infections per seed case after 30 days; (c) probability of elimination of an outbreak starting from a single seed case. Proportion of contacts logged by digital tracing system when both individuals are users of the system is *P*_*d*_ = 90%. Quarantine reduces onward transmission by 50% (blue), by 80% (red), by 80% and with recursive tracing (orange). Isolation of symptomatic cases completely prevents onwards transmission. Error bars show the 95% confidence interval for the mean, calculated from 10,000 independent realisations of the branching process, each run until the outbreak was either eliminated or reached 1,000 cumulative cases.

When quarantine is moderately effective (blue curves in Fig. 3), the addition of digital tracing with high uptake rate (> 75%) and high probability of logging contacts (*P*_*d*_ = 90%) reduced *R*_*eff*_ to around 1.24, mean outbreak size to 20, and increased the probability of extinction to 80%. If quarantine is more effective (reduces transmission by 80%, red curves in Fig. 3), digital tracing can reduce *R*_*eff*_ to approximately 1.12 and increase probability of elimination to 91%. Adding recursive tracing of second-order contacts (orange curves in Fig. 3) provides a relatively small reduction in *R*_*eff*_ to 1.06, although this does increase the probability of elimination from 91% to 95%.

Lower uptake rates (< 75%) result in poorer performance although there is still some noticeable benefit of digital tracing at an uptake rate of around 40%, provided the probability of a close contact being logged is high. If *P*_*d*_ is lower than 90%, performance will deteriorate. However, the results are not as sensitive to *P*_*d*_ as they are to uptake rate *u*, because the requirement for both the index and the secondary case to be users of the system means there is a quadratic dependence on uptake rate, as seen in Figure 3. This means that a 20% reduction in *P*_*d*_ is approximately equivalent to a 10% reduction in uptake rate. We also considered a scenario in which there is no manual contact tracing, except for home contacts which are still assumed to be traced instantly (Figure 4). This could represent a situation where a larger outbreak has exceeded the capacity of the manual contact tracing system, so non-household contacts can only be traced digitally. In this scenario, we assume that quarantine of pre-symptomatic individuals is only moderately (50%) effective and there is no recursive tracing, representing a digital-only contact tracing system without consistent follow-up from trained public health professionals. In this scenario, digital contact tracing makes a larger relative contribution to controlling the spread of COVID-19. However, digital tracing alone is unlikely to be able to contain an outbreak: even with a very high uptake rates (80%) of an effective digital tracing system (*P*_*d*_ = 90%), *R*_*eff*_ is around 1.46. This implies that digital contact tracing would need to be combined with significant population-wide control measures in order to avoid a major epidemic.

**Figure 4:**
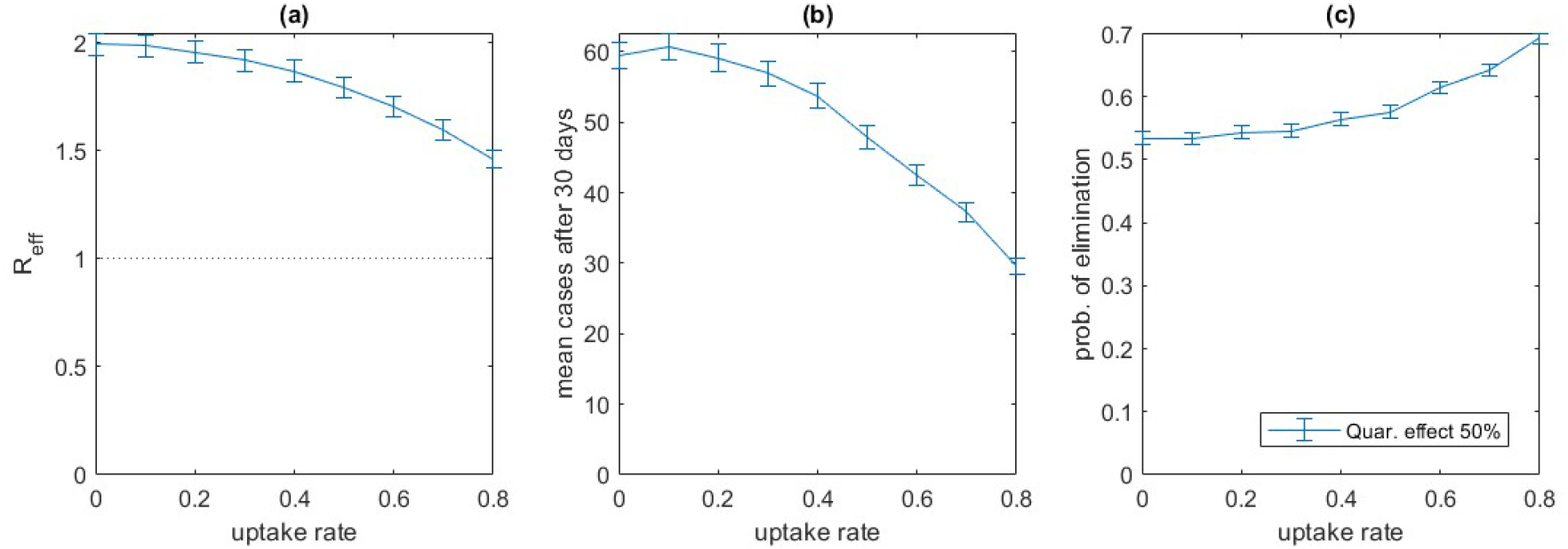
Effectiveness of digital only contact tracing at a range of uptake rates. Effectiveness of a digital tracing system measured by: (a) effective reproduction number *R*_*eff*_ (mean number of secondary infections per case); (b) mean total number of infections per seed case after 30 days; (c) probability of elimination of an outbreak starting from a single seed case.. Home contacts are still traced manually, but other contacts can only be traced digitally. Proportion of contacts logged by digital tracing system when both individuals are users of the system is *P*_*d*_ = 90%. Quarantine reduces onward transmission by 50%. Isolation of symptomatic cases completely prevents onwards transmission. Error bars show the 95% confidence interval for the mean, calculated from 10,000 independent realisations of the branching process, each run until the outbreak was either eliminated or reached 1,000 cumulative cases

Table 2 compares the effectiveness of alternative technological approaches to digital contact tracing, modelled as having different probabilities of recording contacts. Systems based on QR codes alone without proximity detection are likely to perform less well when used as a fully automated tracing system because of the additional steps required for a contact to be recorded: the location needs to have a QR code displayed and both the index case and secondary case need to scan it. This does not diminish the benefit to manual tracing of users scanning QR codes to maintain a record of their movements, but we do not explicitly model this here. Fully decentralised Bluetooth apps are estimated to be less effective than centralised apps at the same level of uptake, because of a reduced likelihood of users reacting to automatic exposure notifications from a decentralised system without follow up from manual contact tracers. A card-based proximity system is estimated to perform similarly to a centralised Bluetooth app, though with a slightly reduced effectiveness because notifications cannot be sent natively and need to be made via a separate system which requires current contact details.

**Table 2.** Comparison of alternative technological approaches to digital contact tracing. Effective reproduction number *R*_*eff*_ of manual contact tracing plus different types of digital tracing system at 40% uptake, 60% uptake and 80% uptake, with highly effective (80%) quarantine and without recursive tracing. A system based on QR codes with no proximity-detection is modelled as logging a low proportion (40%) of contacts, because the location needs to have a QR code displayed, as well as both contacts taking the additional step of scanning the code. Bluetooth apps are modelled as logging a high proportion (90%) of contacts, but in decentralised systems only 50% of users are assumed to quarantine following a notification. A card-based proximity detection system is assumed to have similar detection probability as a Bluetooth app, but only 85% of contacts lead to an instant notification because the card is separate from user’s phones.

| Digital tracing system type | Assumed proportion of contacts lagged | Effective reproduction number $R_{eff}$ | | |
| --- | --- | --- | --- | --- |
|  |  | 40% uptake | 60% uptake | 80% uptake |
| QR code exposure notification | $P_d = 40\%$ of contacts logged | 1.47 | 1.44 | 1.41 |
| Decentralised app-based Bluetooth proximity detection | $P_d = 90\%$ of contacts logged, 50% of notifications result in quarantine | 1.47 | 1.42 | 1.40 |
| Centralised app-based Bluetooth proximity detection | $P_d = 90\%$ of contacts logged | 1.40 | 1.27 | 1.12 |
| Card-based proximity detection | $P_d = 90\%$ of contacts logged, 85% result in an instant notification | 1.41 | 1.34 | 1.22 |

## Discussion

Successful control of COVID-19 is likely to require a range of intervention strategies, including some or all of: moderate population-wide social distancing; widespread use of face coverings; restrictions on large gatherings or other interventions targeting superspreading events; case isolation and household quarantine; manual and digital contact tracing (Ferreti et al. 2020; Hellewell et al., 2020; Kucharski et al., 2020). Establishing trusted relationships with cases and contacts is crucial both to increasing contact tracing coverage and to supporting effective quarantine and isolation. There is still limited evidence on the effectiveness of digital tracing systems (Anglemyer et al., 2020) and reliance on digital tracing alone is unlikely to be sufficient. Digital systems should therefore be seen as an opportunity to improve coverage and/or speed by acting in a complementary role to manual contact tracing. Emphasis should be on how these systems can provide additional useful information to contact tracers in a timely way or fill potential gaps in the manual system. This implies that thorough consultation with public health agencies undertaking contact tracing should be a pre-requisite for an effective digital support system.

In this paper, we modelled the effect of manual contact tracing supported by a digital tracing system with varying levels of uptake and effectiveness. Our results show that manual contact tracing can significantly reduce the spread of COVID-19, but on its own is not sufficient to make the effective reproduction number *R*_*eff*_ less than 1. Manual contact tracing supported by a digital tracing system can further reduce spread, depending on the effectiveness of quarantine of unconfirmed (pre-symptomatic and subclinical) cases. If quarantine is moderately effective (reduces transmission by 50%), a manual plus digital tracing system with very high uptake (> 75%) could reduce *R*_*eff*_ to around 1.2. If quarantine is highly effective (reduces transmission by 80%), *R*_*eff*_ can be reduced to approximately 1.1, which could be sufficient to contain the majority of small outbreaks. However, if the uptake rate is less than around 75%, the reduction in *R*_*eff*_ is not sufficient to contain an outbreak without additional measures.

Our results suggest that the additional benefit from recursive tracing of second-order contacts is relatively small. In the case of a small outbreak, this may be worthwhile as it could increase the probability of elimination from 90% to 95% provided uptake is high and quarantine is effective. However, given that it is likely more difficult to effectively quarantine second-order contacts, and the number of uninfected people being quarantined would be much higher (Firth et al., 2020), it may be more beneficial to focus on effective quarantine of first-order contacts than attempting to locate second-order contacts.

In our model, untraced clinical cases take 6.8 days on average to get tested after onset of symptoms. The aim of contact tracing is to find close contacts of confirmed cases and therefore enable early quarantine or isolation. However, it is important to recognise that reducing the time between symptom onset and isolation can bring significant benefits, even in the absence of contact tracing. This implies that enhancing public awareness of COVID-19 symptoms and the need to get tested quickly, ensuring equitable access to healthcare, and maintaining rapid testing capacity are equally important as investment in contract tracing.

We have not considered the consequences of false positives from digital contact tracing systems, i.e. people who are recorded as potential contacts by the digital system but have not in fact been at risk of exposure to COVID-19. For countries that have reduced cases to very low numbers and where the primary goal is to achieve or maintain elimination of community transmission, we assume that the number of false positives is not a primary consideration.

If case numbers exceed the capacity of the manual contact tracing system, its performance will rapidly deteriorate. Under this scenario, digital tracing can make a significant contribution to slowing the growth of the outbreak, but if it became the dominant form of tracing it is likely to be insufficient to reduce *R*_*eff*_ under 1. This implies that population-wide control measures would likely be needed to prevent a major epidemic. A low false positive rate is a more important consideration in this situation.

Our model allowed for age-specific mixing patterns in home, work, school, and casual contacts, and for heterogeneity and homophily in individual contact rates. However, the model ignores other sources of heterogeneity, for example in types of workplace. Some workplaces will be much more amenable to rapid contact tracing than others, for example where employees tend to work in a consistent physical location each day. This could be modelled via individual heterogeneity and homophily in contact tracing probabilities as well as contact rates, however the mean tracing probability is likely to remain the most important parameter.

We have taken a technology-agnostic approach to modelling digital contact tracing. Among the most important parameters for any digital tracing system are the uptake rate (number of people using the system) and the probability of close contacts being logged. Our results show that to contain an outbreak, a well-functioning manual contact tracing system needs to be combined with a digital tracing system that should ideally have at least 75% uptake and record 90% of close contacts. Different systems may have different uptake rates, for example due to usability and privacy issues, so a careful study of expected uptake rates is critical to choosing the best system. The proportion of contacts logged will be affected by the usability of the system and by individual behaviour. For example, contacts will be missed in situations where a user forgets or loses their phone or card, has the phone switched off or a flat battery, or has Bluetooth deactivated. Systems that rely solely on QR code scanning for exposure notification are likely to perform less well than systems with proximity-based detection at the same level of uptake. This is because successful tracing requires the location to have a compatible QR code displayed, both individuals to have the app installed and to take the additional step of scanning the code. There may be benefits to a QR code app in keeping a record of movements or digital diary to assist manual contact tracers to identify contacts who would otherwise be missed. We did not explicitly consider this, though it could be modelled via an increase in the manual tracing probabilities for relevant settings.

## Data Availability

No new data is presented in this manuscript

## Acknowledgements

The authors acknowledge the support of StatsNZ, ESR, and the Ministry of Health in supplying data in support of this work. The authors are grateful to Andrew Chen, Matt Parry and Philippa Yasbek for discussions about digital contact tracing systems and feedback on an earlier version of this paper. This work was funded by the Ministry of Business, Innovation and Employment and Te Pūnaha Matatini, New Zealand’s Centre of Research Excellence in complex systems.

