## Supplementary Material for "Potential reduction in transmission of COVID-19 by digital contact tracing systems: a modelling study"

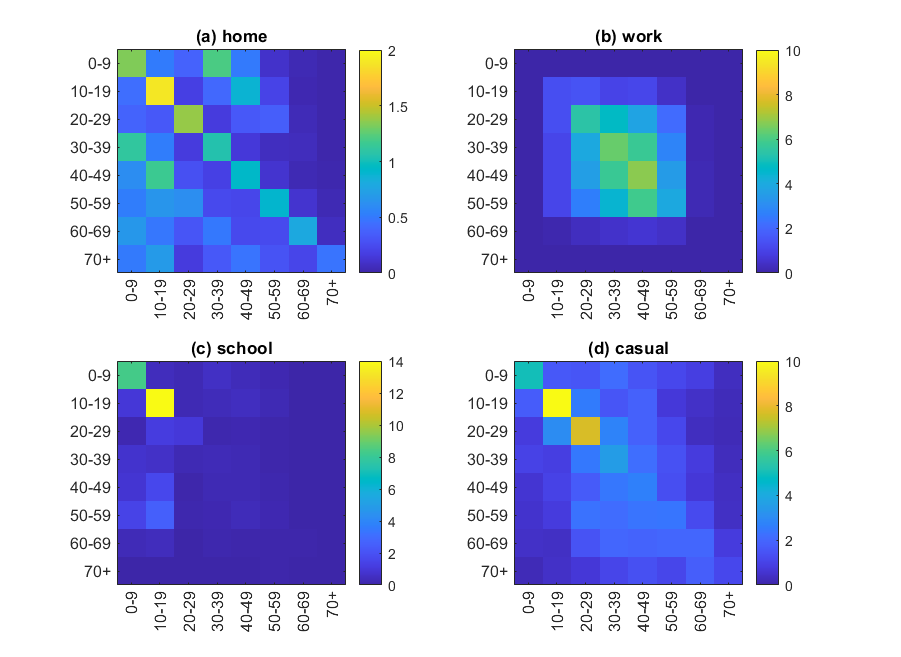

**Supplementary Figure S1.** Contact matrices in 10-year age bands derived from results of Prem et al. (2017) using the number of daily contacts for New Zealand combined into 10 year age bands. Each matrix shows the average number of home, work, school or casual contacts that an individual in age group $k$ has with an individual in age group $l$ over the course of their infectious period. The number of home contacts are taken directly from the results of Prem et al. (2017) for New Zealand. The number of work, school and casual matrices are scaled up by a factor of 3 from the number of daily work, school, and other contacts in Prem et al. (2017) to allow for an infectious period longer than 1 day. The factor of 3 was chosen to give a basic reproduction number of $R_{0}=2.6$ in the absence of any control measures.

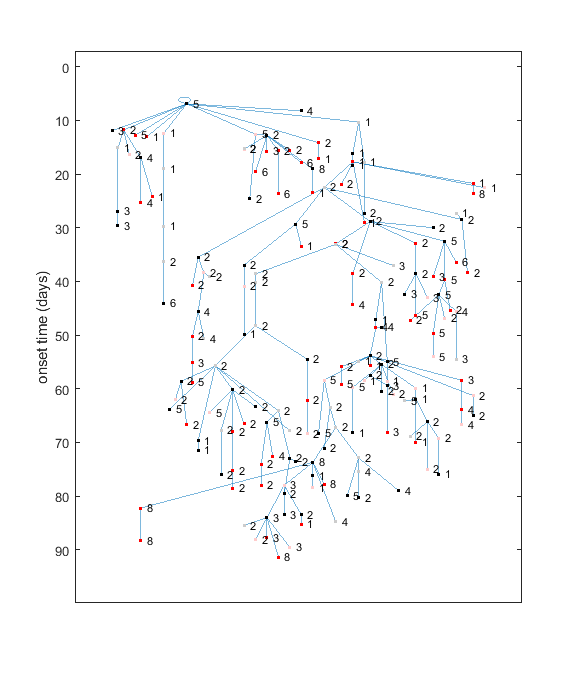

**Supplementary Figure S2.** Example branching process simulation starting from a single infected seed case. Cases are represented as nodes with the vertical coordinate corresponding to time of symptom onset. Transmission routes are blue lines. Node labels indicate age group (1 = 0-10 years, 2 = 10-20 years, etc.), red indicates traced contacts, black indicates untraced, grey/pink indicates subclinical individuals.
